# Mentalizing abilities mediate the impact of the low-level social cognitive processes on negative symptoms

**DOI:** 10.1101/2022.01.26.22269892

**Authors:** Ł. Okruszek, M. Chrustowicz, M. Jarkiewicz, M. Krawczyk, V. Manera, A. Piejka, A. Schudy, M. Wiśniewska, A. Wysokiński

## Abstract

Social cognitive deficits are currently considered as one of the main predictors of clinical symptoms and functional outcome in patients with schizophrenia. Multiple studies have suggested that two-factor solution (low-level *vs*. high-level) best describes the structure of social cognitive processes in patients. While higher-order processes have been repeatedly linked to negative symptoms, no such association was found for lower-level processes. Thus, the aim of the current study is to examine whether the association between low-level socio-perceptual processes and symptoms in patients with schizophrenia is mediated by higher-order socio-inferential abilities. One hundred thirty-nine patients have completed basic communicative interactions processing (CID-12) and mind reading (Reading the Mind in the Eyes task) tasks. In line with our hypothesis, we have observed full mediation of the effects of basic social perception abilities on negative symptoms via mentalizing abilities in patients. This effect suggests that, similarly as in the case of positive symptoms, a hierarchical nature of social cognitive processes should be considered while investigating negative symptoms of schizophrenia.

## Introduction

Investigation of social cognitive deficits is currently recognized as one of the main areas of translational research in psychiatry, in particular in the field of the schizophrenia research (Vaskinn and Horan 2020). While the initial interest was focused on general neurocognition and nonsocial cognitive deficits in patients, the social cognitive domain has been soon added to the MATRICS Consensus Cognitive Battery, a ‘gold standard’ method for assessment of cognition in patients with schizophrenia (SCZ) (Nuechterlein et al. 2008). Since then, large-scale initiatives have also focused on creating psychometrically sound methods for the assessment a wide range of social cognitive processes in patients (Pinkham et al. 2018). Moreover, multiple studies have supported the notion that social cognition is a better predictor of the community functioning than nonsocial cognition (Fett et al. 2011) and that it may be a mediator of the relationship between nonsocial cognition and functional outcome in SCZ (Schmidt et al. 2011). This effect however has been attributed to social cognitive abilities being more directly linked to skills that are crucial for everyday functioning than to the general nonsocial cognitive capacity. Nevertheless, negative symptoms have been recently conceptualized as ‘real-world’ consequences of social cognitive deficits (Pelletier-Baldelli and Holt 2020). This way, a pathway linking cognitive deficits with functional outcome via negative symptoms, which are a strong predictor of functional outcome in patients (Ventura et al. 2009), can also be considered.

The interpretation of communicative interactions is a complex social cognitive ability that requires dynamic cooperation of both automatic and controlled processes, and is linked to increased activity and functional connectivity of the main social brain networks (Quadflieg and Koldewyn 2017). Exploring the mechanisms associated with communicative interaction processing may be crucial for understanding problems that are encountered by SCZ during their everyday functioning. We have presented an initial evidence for disturbed recognition of communicative interactions from point-light motion in schizophrenia on the basis of the preliminary sample of eighteen SCZ and eighteen healthy controls (Okruszek et al. 2015). As the initial analyses suggested that these effects may be dragged by the misclassification of dyadic individual actions as communicative interactions, we tended to interpret these initial findings as an evidence for “overmentalizing” (Frith 2004) in SCZ. This notion was also supported by the relationship between interpretation of dyadic actions and ability to attribute complex emotional states on the basis of the facial display observed in patients. However, our subsequent study which measured both reflexive and reflective social cognitive processes associated with communicative interaction processing in patients and controls has found an overall decrease in recognition of explicitly presented communicative interactions in a sample of forty-six patients and forty controls, while reflexive social cognitive processes were at least partially preserved (Okruszek et al. 2018a). Yet, in line with the first study we have found a correlation between mentalizing ability, as measured by the Reading Mind in the Eyes task (RMET) and recognition of dyadic individual actions in the second study.

While the overall functional organization of social cognitive processes in SCZ is still subject of debate, studies in the field present evidence for one-up to four-factor structure social cognition. Currently, the most prevalent pattern of findings favors the two factor model which distinguishes low-level social cognitive abilities associated with basic social stimuli processing and higher-level abilities associated with inferring complex mental states (Etchepare and Prouteau 2018). Moreover, the studies that investigated the association between social cognitive factors and symptoms in patients found associations between higher-order social cognitive abilities and symptoms, while no such relationship was found for low-level social cognitive abilities (Mehta et al. 2014a; Oliver et al. 2019; Riedel et al. 2020). At the same time, given the strong intercorrelation between lower- and higher-level factors (e.g., Pearson’s r=.35 in Mehta et al., 2014; r=.50 in Riedel et al., 2020), one may hypothesize that the association between low-level social cognitive processes and symptoms in patients is mediated by higher order-social cognitive processes.

Unfortunately, the power sample of none of the studies on communicative interaction processing in schizophrenia was suitable to reliably test for mediation effects, assuming medium to large association strength observed across two studies (Fritz and Mackinnon 2007). Thus, the aim of the current study is to test the hypothesis that the association between a low-level communicative interaction processing and symptoms observed in SCZ is mediated by the mentalizing abilities.

## Methods

### Participants

The clinical sample for the current study was obtained merging the participants originally reported in Okruszek et al., (2018a) and an additional sample of participants recruited as a part of a larger study that investigated EEG-fMRI markers of social perception in schizophrenia (Polish National Centre of Science 2016/23/D/HS6/02947 study, PI: Łukasz Okruszek). Overall, one hundred thirty-nine participants diagnosed with schizophrenia according to ICD-10 criteria were recruited from the Department of Old Age Psychiatry and Psychotic Disorders, Medical University of Łódź (n=45) and Institute of Psychiatry and Neurology in Warsaw (n=94). Adult (>18 years old) patients with no history of other psychiatric or neurological comorbidities or intellectual disability were included in the study. The protocol of each study was approved by the respective bioethics committee (either Medical University of Lodz Bioethics Committee or Ethics Committee at the Institute of Psychology, Polish Academy of Sciences). Severity of clinical symptoms was examined in patients by a trained psychiatrist using Positive and Negative Syndrome Scale (Kay et al. 1987). The clinical domains were calculated in line with PHAMOUS consortium, which resulted in four clinical indicators: positive, negative, affective and cognitive symptoms (Chen et al. 2020) (Table 1). For the current analysis only individuals with no missing data for any variables of interest (CID-12, RMET, and, in case of patients, PANSS) were included in the analyses. Healthy participants with no known history of psychiatric or neurological disorders were recruited via online advertisements and included in the study as a control group. Controls were matched head-to-head to patients with regard to age (SCZ: 33.52+/-8.23; HC: 33.80+/-6.78 t(276)=.310 p=.76) and sex (45F/94M in each group).

**Table 1.** Basic descriptive statistics for PHAMOUS symptom domains in patients.

|  | Range | Mean | Standard deviation |
| --- | --- | --- | --- |
| PHAMOUS<br>POSITIVE | 4-22 | 9.74 | 4.69 |
| PHAMOUS<br>NEGATIVE | 7 - 36 | 15.62 | 6.35 |
| PHAMOUS<br>AFFECTIVE | 7-37 | 16.57 | 6.48 |
| PHAMOUS<br>COGNITIVE | 12-41 | 23.25 | 7.08 |

### Procedure

During the behavioral assessment session participant completed Communicative Interaction Database explicit recognition task (Manera et al. 2015) and Reading the Mind in the Eyes task (Calder et al. 2002) as proxies of either lower-level social perception abilities and higher level social inference abilities. In each study, CID-12 and RMET were performed as a part of a broader assessment battery, however the selection of the tasks for the current study was limited by the setup of the original study which focused mostly on psychophysics tasks (Okruszek et al. 2018a). Detailed description of CID-12 and RMET may be found below.

### CID-12

We have utilized a short version of the Communicative Interaction Database explicit recognition task, which consisted of consisted of 12 videos of point-light actions depicting two point-light agents selected from the Manera et al.’s (Manera et al. 2010) database. The presented actions included six communicative actions (COM: “pick up”, “choose one object”, “put object down”, “look at the ceiling”, “get down”, “sit down”) and six individual actions (IND: “jump”, “look at the foot”, “sneeze”, “lateral step”, “drink”, “turn”). Each action was presented twice. Upon the second presentation of each video participants were asked: (i) to decide whether the two agents were communicating versus acting independently of each other; and (ii) to select the best description of the action from five alternatives (Manera et al. 2015). Each set of alternatives consisted of (i) a correct description of actions of both agents, (ii) two incorrect answers which have been created by replacing agent A’s action with communicative action and (iii) two incorrect answers which have been created by replacing agent A’s action with individual action. No time limit was imposed on participants while giving answers. Score from each sub-task (detection of communicative interactions; identification of specific actions) for each type of the stimuli (communicative interactions; dyadic individual actions) are subjected to further analysis.

### Reading Mind in the Eyes Test

Mentalizing abilities were examined using the Polish version of the RMET task (Calder et al. 2002). During the task the participant is presented with thirty-six black-and-white photographs of eye-region of the face taken from photographs of different actors (19) and actresses (17). Each vignette is surrounded by four words describing various complex mental states (e.g., serious; ashamed; alarmed; bewildered). The participant is asked to choose the best description of what the person in the photograph may be feeling or thinking.

### Statistical analysis

Data analyses were performed using Statistical Package for the Social Sciences (SPSS 27). A repeated-measures ANOVA with stimulus type (communicative *vs*. individual) and task (detection of communicative interactions *vs*. identification of specific actions) as within subject factors and group (SCZ *vs*. HC) as between-subject factors was used to examine the CID-12 scores. Next, the association between CID-12 scores and RMET was examined in each group separately using Pearson correlations. In patients, the association between CID-12 scores and PHAMOUS domains was examined using Pearson correlations. Finally, a mediation effects between CID-12 score (averaged across stimuli and conditions), RMET score and PHAMOUS Negative was examined using PROCESS 4.0 by examining whether percentile bootstrap confidence intervals contained 0.

## Results

### CID-12

Between-group differences: A main effect of the task (F(1, 276)=141.8; p<.001; η^2^_p_=.34; 95% CI=[.25; .42]) was found, with higher accuracy for detection of communicative interactions *vs*. individual actions (4.87+/-0.97) compared to identification of specific actions (4.08+/-1.12). Moreover, an interaction between stimulus type and task was found, with higher accuracy during detection compared to identification task for communicative actions and reverse pattern for individual actions (F(1, 276)=32.3; p<.001; η^2^_p_=.11; 95% CI=[.05; .18]). Finally, patients performed worse in CID-12 task compared to healthy controls (F(1, 276)= 71.1; p<.001; η^2^_p_=.21; 95% CI=[.13; .28]). No other significant effects were found. Results of the task are visualized in the Figure 1.

**Figure 1.**
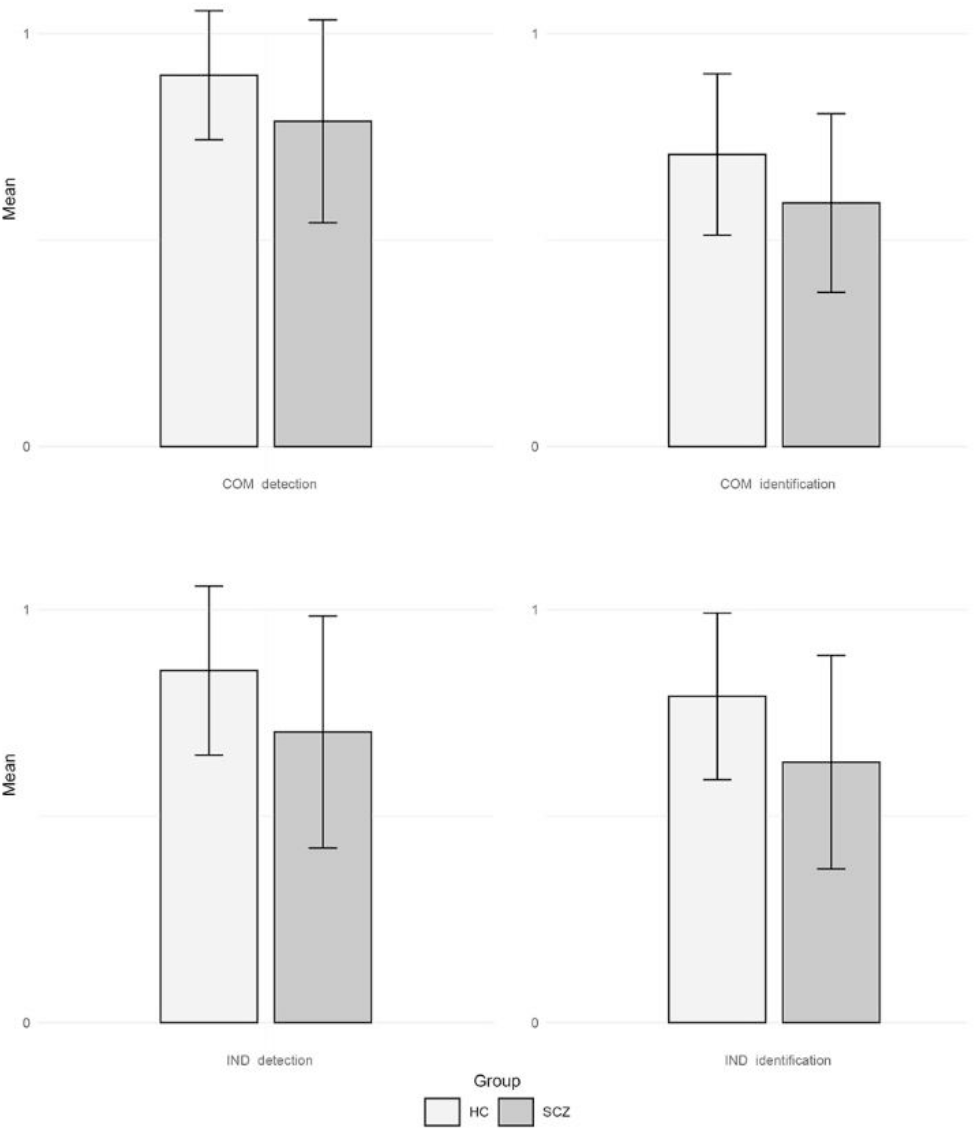
Results from the CID-12 task: on the left - percentage of correct responses for categorization as communicative or individual, on the right: percentage of correct responses for selecting the description of the actions from five alternatives). Upper panel – percentage of correct responses for trials depicting communicative interactions (COM), lower panel - percentage of correct responses for trials depicting dyadic individual actions (IND). HC – Healthy Controls; SCZ – Patients with schizophrenia.

### RMET

Controls (25.7+/-3.9) outperformed patients (23.2+/-4.9; F(1, 271) =22.0; p<.001; η^2^_p_=.08; 95% CI=[.03; .14]).

### Correlations

Zero order correlations between variables are shown in Table 2.

**Table 2.** Zero order correlations between the variables of interest. CID – Communicative Interaction Database, RMET – Reading the Mind in the Eyes Test, HC – Healthy Controls, SCZ – Patients with schizophrenia

|  | RMET HC | RMET SCZ | PHAMOUS<br>NEGATIVE | PHAMOUS<br>POSITIVE | PHAMOUS<br>AFFECTIVE | PHAMOUS<br>COGNITIVE |
| --- | --- | --- | --- | --- | --- | --- |
| CID 1 COM | .10 | .13 | .03 | .13 | .06 | .11 |
| CID 1 IND | -.02 | .11 | -.13 | -.17* | -.15 | -.11 |
| CID 1 SUM | .06 | .17* | -.08 | -.05 | -.08 | -.01 |
| CID 2 COM | .29*** | .26** | -.20* | -.31*** | -.13 | -.36*** |
| CID 2 IND | .18* | .37*** | -.11 | -.14 | -.18* | -.11 |
| CID 2 SUM | .30*** | .40*** | -.19* | -.27** | -.19* | -.28*** |
| CID total | .24** | .36*** | -.17* | -.20* | -.17* | -.18* |
| RMET | - | - | -.22* | -.13 | -.05 | -.21* |

### Mediation analysis

The CID-12 explained 13% of RMET variability (F(1, 133)=20.2; p<.001; β=.366). The model with RMET and CID-12 explained 6% of negative symptoms (F(2, 132)=4.0; p<.05), with RMET being the only significant predictor of negative symptoms (β=-.184; p<.05). However, a bootstrapping analysis revealed a significant indirect mediation effect of CID-12 on negative symptoms via mentalizing (β=-.067; CI=[-.142, -.004]) (Figure 2). For no other group of symptoms similar effects emerged. Importantly, the sensitivity of the observed results was ascertained, as no significant indirect effect was found in analysis with RMET as IV and CID-12 as mediator (β=-.035; CI=[-.103, .022]).

**Figure 2.**
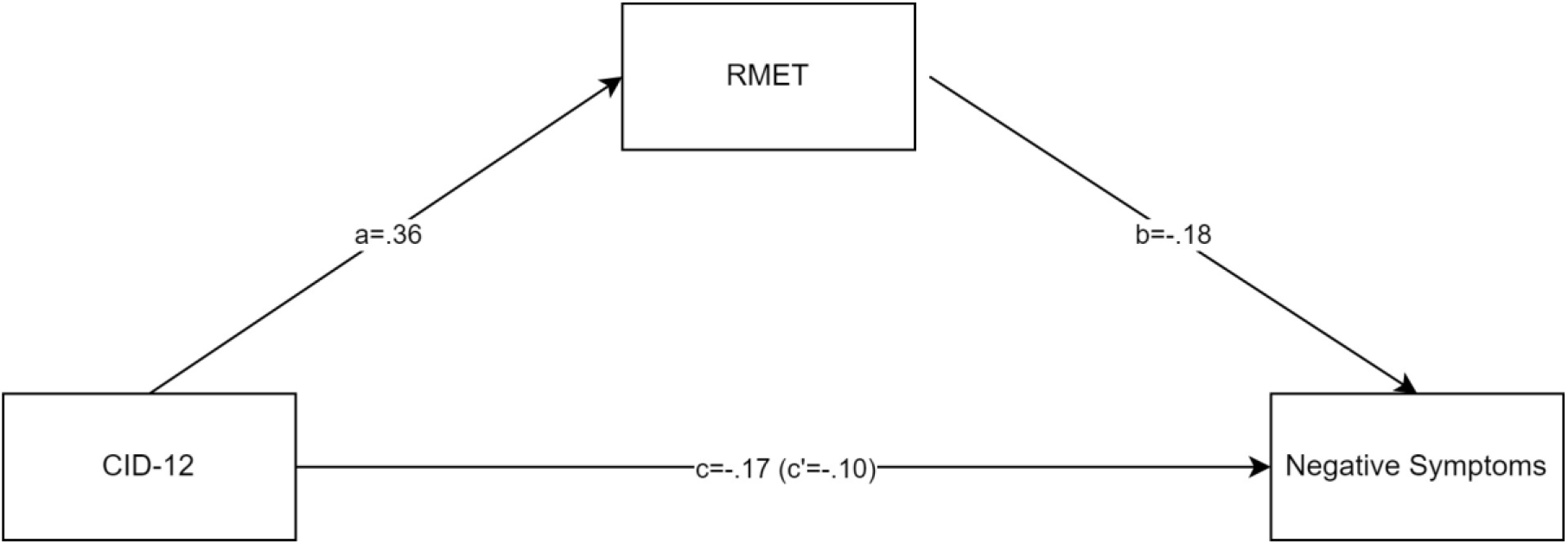
Mediation of the basic social perception (Communicative Interaction Database-12; CID-12) on negative symptoms via mentalizing abilities (Reading the Mind in the Eyes Test; RMET).

## Discussion

The aim of the current mega-analysis was to investigate the ability to process communicative interactions from point-light motion in a well-powered sample of SCZ and a well-matched group of healthy controls. Furthermore, we aimed to test whether associations between low-level processing of communicative interactions and symptoms is mediated by the higher-order mentalizing abilities in patients.

Firstly, we have found overall worse performance in communicative interactions processing task in SCZ compared to healthy controls. This effect was not modulated by the type of task or the type of the stimuli presented during the task, which provides support for the notion of generalized nature of deficit in this area in this clinical group. This finding contradicts our initial tentative report that patients present similar ability to detect communicative interactions as healthy controls in CID-12 (Okruszek et al. 2015). However, in the current study, the interaction between task and stimuli was observed and the highest accuracy was observed in all participants for the classification of the stimuli as communicative *vs*. individual during the detection task in CID-12. Given the possibly underpowered sample of the original study, the lack of observed differences for the classification of the communicative vignettes may be attributed to the psychometric confound (Kang and MacDonald 2010). Interestingly, the average effect size observed for the communicative interactions processing task in the current study was larger (d=1.01) than for mentalizing task (d=0.57). In general, the meta-analytic reviews suggest the opposite pattern of results: Savla et al. (Savla et al. 2013) observed large effect sizes (g=0.96) across fifty studies that compared accuracy on Theory of Mind tasks in 1760 SCZ and 1536 controls (including eleven studies based on RMET). At the same time, we have previously reported an overall moderate-to-large effect (d=0.66) for biological motion processing in a meta-analysis of 14 studies with 571 patients and 482 controls (Okruszek and Pilecka 2017).

Secondly, we have found significant correlation between lower-level processing of communicative interactions from point-light motion and the accuracy on the Reading Mind in Eyes task. The association between lower and higher-order social cognitive abilities was observed to be stronger in patients compared to controls. Additionally, while mentalizing abilities were associated with interpretation of specific actions in both groups, in patients a weak association between RMET performance and detection of communicative interactions was also found. These finding are congruent with previous suggestions that social cognitive skills are less differentiated in patients than in healthy individuals, e.g. Buck et al. (Buck et al. 2016) found two-factor solution (low-level *vs*. high-level processes) in healthy controls, but only a single factor solution in patients.

Finally, in line with the main hypothesis of the study we have found both direct and indirect association of communicative interaction processing with symptoms in patients. Total CID-12 score has shown negative correlations with all four domains of symptoms included in the PHAMOUS factoring. Furthermore, as mentalizing skills were negatively associated with negative and cognitive symptoms in patients, indirect paths linking communicative interactions processing capacity with these domains via mentalizing abilities were tested. Full mediation effect was found, with significant indirect path linking basic social perception with negative symptoms via ToM in patients. This pattern suggests that negative symptoms in patients are linked with the inability to overcome misinterpretation of basic social cues via higher order mental state reading, rather than with basic social perception deficits *per se*. Thus, “two-factor” framework may be proposed, i.e., some negative symptoms (e.g., social withdrawal) in patients may be elicited by (1) lower-level socio-perceptual deficits which are not counteracted and may be further augmented by (2) deficient higher-order socio-inferential abilities. In addition, while a large body of research and theoretical discussion has been directed toward explaining positive symptoms in the light of the hierarchical Bayesian nature of perceptual and inference mechanisms (Fletcher and Frith 2009), such hierarchical conceptualizations are largely absent with regard to other groups of symptoms. Furthermore, practical implications of the current finding can be pointed out – a recent review has included limited effectiveness and generalizability of social cognitive interventions into other domains of functioning among unresolved issues and challenges in the field of research on social cognition and schizophrenia (Vaskinn and Horan 2020). Recent network meta-analysis has shown that while all types of treatments included in the analysis significantly improve social perception in patients, only broad-based social cognitive training improves ToM processes in patients (Nijman et al. 2020). Hence, given the trajectory of processes linking basic social cognitive processes with negative symptoms observed in the current study, broad and multifaceted interventions, may be more likely to achieve clinical improvements in patients compared to highly targeted interventions that address limited set of social cognitive processes.

The current study investigated trajectories linking communicative interactions processing, mentalizing and negative symptoms in SCZ. While it had been informed by our extensive previous research in this area and included a well-powered sample of participants and well-matched healthy controls, still some limitations of the current study should be taken into consideration. First of all, while our hypotheses were informed by studies which investigated social cognitive factors in schizophrenia, in the current study lower- and higher-order processes were indicated by a single task score. Although we advocate for using broad battery of measures with known psychometric abilities in clinical and non-clinical samples (e.g. (Okruszek et al. 2021), the design of the current study was bounded by the set of tests included in our previous study (Okruszek et al. 2018a), thus we could not use neurocognitive or social cognitive variables available for the new subset of participants.

Furthermore, while RMET is predominantly conceptualized as a ToM task, it heavy relies on the perceptual processes. Oppositely, while the CID-12 is a low-level socio-perceptual task, it may involve some degree of intention attribution (Centelles et al. 2011; Okruszek et al. 2018b). Thus, neither of the two tasks used in the current study can be considered as a “pure” measure of lower- or higher-order social cognitive processes. Furthermore, it has been shown that the trajectory linking social cognition and negative symptoms may be further expanded, as it has been shown that negative symptoms mediate the influence of Theory of Mind on functional outcome variables in SCZ (Mehta et al. 2014b). Along these lines, future studies should test sequential mediation effects to elucidate the indirect role of low-level social cognitive processes in functional outcome in patients.

## Data Availability

All data produced in the present study are available upon reasonable request to the authors

## Acknowledgments

This work was supported by the National Science Centre, Poland (Grant No: 2016/23/D/HS6/02947).

